# Treatment for radiographically active, sputum culture-negative pulmonary tuberculosis: a systematic review and meta-analysis

**DOI:** 10.1101/2023.01.27.23285085

**Authors:** Adam Thorburn Gray, Liana Macpherson, Ffion Carlin, Bianca Sossen, Alexandra S Richards, Sandra V Kik, Rein M G J Houben, Peter MacPherson, Matteo Quartagno, Ewelina Rogozińska, Hanif Esmail

## Abstract

**INTRODUCTION:** People with radiographic evidence for pulmonary tuberculosis (TB), but negative sputum cultures, have increased risk of developing culture-positive TB. Recent expansion of X-ray screening is leading to increased identification of this group. We set out to synthesise the evidence for treatment to prevent progression to culture-positive disease.

**METHODS:** We searched for prospective trials evaluating the efficacy of TB regimens against placebo, observation, or alternative regimens, for the treatment of adults and children with radiographic evidence of TB but culture-negative respiratory samples. Databases were searched up to 18 Oct 2022. Study quality was assessed using ROB 2.0 and ROBINS-I. The primary outcome was progression to culture-positive TB. Meta-analysis with a random effects model was conducted to estimate pooled efficacy.

**RESULTS:** We included 13 trials (32,568 individuals) conducted between 1955 and 2018. Radiographic and bacteriological criteria for inclusion varied. 19.1% to 57.9% of participants with active x-ray changes and no treatment progressed to culture-positive disease. Progression was reduced with any treatment (6 studies, risk ratio [RR] 0.27, 95%CI 0.13-0.56); multi-drug TB treatment (RR 0.11, 95%CI 0.05 - 0.23), was significantly more effective than isoniazid treatment (RR 0.63, 95%CI 0.35-1.13) (p=0.0002).

**DISCUSSION:** Multi-drug regimens were associated with significantly reduced risk of progression to TB disease for individuals with radiographically apparent, but culture-negative TB. However, most studies were old, conducted prior to the HIV epidemic and with outdated regimens. New clinical trials are required to identify the optimal treatment approach.

**STUDY REGISTRATION:** CRD42021248486

## INTRODUCTION

Globally in 2021, 2 million (37%) of the 5.3 million people notified with pulmonary tuberculosis (TB) were not bacteriologically confirmed. This percentage has remained unchanged over recent years[1] and is also found in high resource settings, such as the UK (39.3%).[2] Lack of bacteriological confirmation may be due to technical factors including sample quality, laboratory processing, diagnostic test sensitivity,[3] or clinical factors as some individuals have paucibacillary TB disease (e.g., children, people living with HIV, extra-pulmonary disease). For these reasons, inability to detect *Mycobacterium tuberculosis* does not exclude active disease and in some circumstances radiology, symptomatology, or histology, potentially combined with exposure history, are used to inform treatment decisions.

Active pulmonary TB is usually confirmed by a sputum bacteriological test (e.g., smear, molecular test, culture). However, disease pathology and infectiousness can precede symptom development; and these individuals with incipient or subclinical disease may be detectable radiographically.[4] This has long been recognised and in the mid-twentieth century mass chest X-ray (CXR) screening for TB was widely implemented. After the 1970s the cost effectiveness of the approach was questioned, especially in countries experiencing a rapid decline in incidence, and the practice was largely abandoned.[4] However, increasingly affordable and accessible technology (e.g., digital X-ray, computer-aided-detection software) has led to a re-expansion of CXR-based screening, and the potential utility in increasing case detection has been re-emphasised in the updated WHO TB screening guidance.[6] However, the optimal management for individuals with CXR abnormalities but negative confirmatory sputum tests is unknown. Importantly, for those who don’t receive treatment, the risk of progression from bacteriologically negative to positive TB is approximately 10% per year.[7]

Over the last 30 years management of TB has remained rooted in a binary approach: treat for so-called ‘active’ TB disease or ‘latent’ TB infection.[8] For drug-susceptible active TB in adults (excluding central nervous system disease) a one-size-fits-all strategy prevails irrespective of bacillary burden.[6] The standard 6-month treatment of rifampicin and isoniazid supplemented with pyrazinamide and ethambutol for the first two months was developed in trials designed to identify successful treatments for smear positive disease.[9] Hence, treatment duration and composition are driven by the requirements of the most extensive, multibacillary forms of disease, potentially over treating more paucibacillary states.[8] Recognising the false dichotomy of binary disease management, the recent SHINE trial has shown that minimal, largely bacteriologically negative, disease in children can be treated with a four-month regimen.[10]

The optimal regimen and duration for bacteriologically negative, active pulmonary TB in adults is not known. These patients are less likely to be symptomatic at time of diagnosis and may be less willing to tolerate the duration, pill burden, and drug toxicity of the standard 6-month regimen.[11] Shorter, less toxic regimens could reduce adverse events, increase adherence, and reduce individual and programmatic costs, but clinical trials are needed to determine effectiveness. We sought to systematically identify evidence from clinical trials that compared the effectiveness of anti-tuberculous regimens to placebo, observation, or alternative regimens, on disease progression in people with bacteriologically negative, but radiographically apparent pulmonary TB.

## METHODS

### Search strategy and selection criteria

We searched EMBASE (Ovid from 1947), MEDLINE (Ovid from 1946), the Cochrane Infectious Diseases Group specialised register, and Web of Science (from 1900) with no language restrictions. Eligible studies were randomised and non-randomised prospective trials comparing treatment against placebo, observation, or alternative treatment, for the management of children and adults with suspected pulmonary TB based on CXR findings, but with negative respiratory tests (culture or molecular test); the search strategy is available (Supplementary Material). Initial searches were performed on 17^th^ March 2021 and updated on 18^th^ October 2022; abstracts and eligible full manuscripts were independently screened by two of LM, FC, or AG, with discrepancies resolved by HE. References of included articles were reviewed for additional relevant articles. This systematic review and meta-analysis are reported in accordance with the Preferred Reporting Items for Systematic Reviews and Meta-Analyses (PRISMA).[12]

### Data extraction and quality assessment

Data were independently extracted by two of LM, FC, or AG, into a piloted database to inform a primary outcome of progression to bacteriologically confirmed pulmonary TB, defined as at least one positive smear, culture, or molecular test on sputum, laryngeal swab, or gastric lavage during follow up. Further data were extracted where available to inform secondary outcomes including clinical or radiographic evidence of progressive active TB at multiple time-points (0, 6, 12, 24, 48, 60 months); adverse events; drug resistance; treatment adherence; and loss to follow-up (Supplementary Table S1). Data on country-level TB incidence were extracted to inform baseline risk of TB disease if available. Two reviewers independently assessed risk of bias of included studies using recommended tools for randomised (RoB 2.0)[13] and non-randomised trials (ROBINS-I).[14]

### Statistical analysis

Clinical and epidemiological characteristics were summarised, including the number and percentage of participants excluded at baseline due to positive cultures and the duration of follow-up. To investigate the effect of treatment regimens on progression to bacteriologically confirmed TB, we pooled data from comparable trials (active radiographic changes and use of an inactive comparator) using a random-effects model with restricted maximum likelihood (REML) estimation of variance method. The effects are reported as pooled unadjusted relative risks (RR) and number-needed-to-treat (NNT, i.e. the number of participants that would be required to be treated to avert one progression to active pulmonary TB).[15] For studies or cohorts with zero incident cases over the follow-up period, a fixed value was added. Heterogeneity was assessed by inspecting forest plots and by calculating the I^2^ statistic and explored using subgroup analyses: treatment regimen composition (multi-drug regimens vs simple isoniazid-based regimens; stratification determined after qualitative review of included studies); case finding methodology (active case-finding [ACF] vs passive case-finding [PCF]); number of baseline sputa samples (fewer than three vs three or more); HIV prevalence in participants (<1% vs >1%); and severity of baseline radiographic abnormality (single lobar disease vs more extensive disease). A funnel plot was constructed to assess small study effects and potential for publication bias. Sensitivity analyses were conducted using studies with only low or some risk of bias (as defined by bias assessment tools), using all TB diagnoses (all individuals diagnosed with TB disease with or without bacteriological confirmation) for the primary outcome, and using per protocol results (assessing those that completed study follow up). We planned to conduct a network meta-analysis if the data supported this. Analyses were performed using R (Version 4.2.2, R Foundation for Statistical Computing, Vienna) using the metafor package,[16] and Stata (version 17.0, Statacorp, College Station, Texas, USA).

## RESULTS

We identified 9733 unique publications for title and abstract screening of which 41 met criteria for full text review. Of these 18 were included, with one additional publication identified through reference screening, resulting in 19 manuscripts relating to 13 studies (Figure 1 and Supplementary Material). Studies excluded at full text review are listed with rationale for exclusion (Supplementary Table S2).

**Figure 1:**
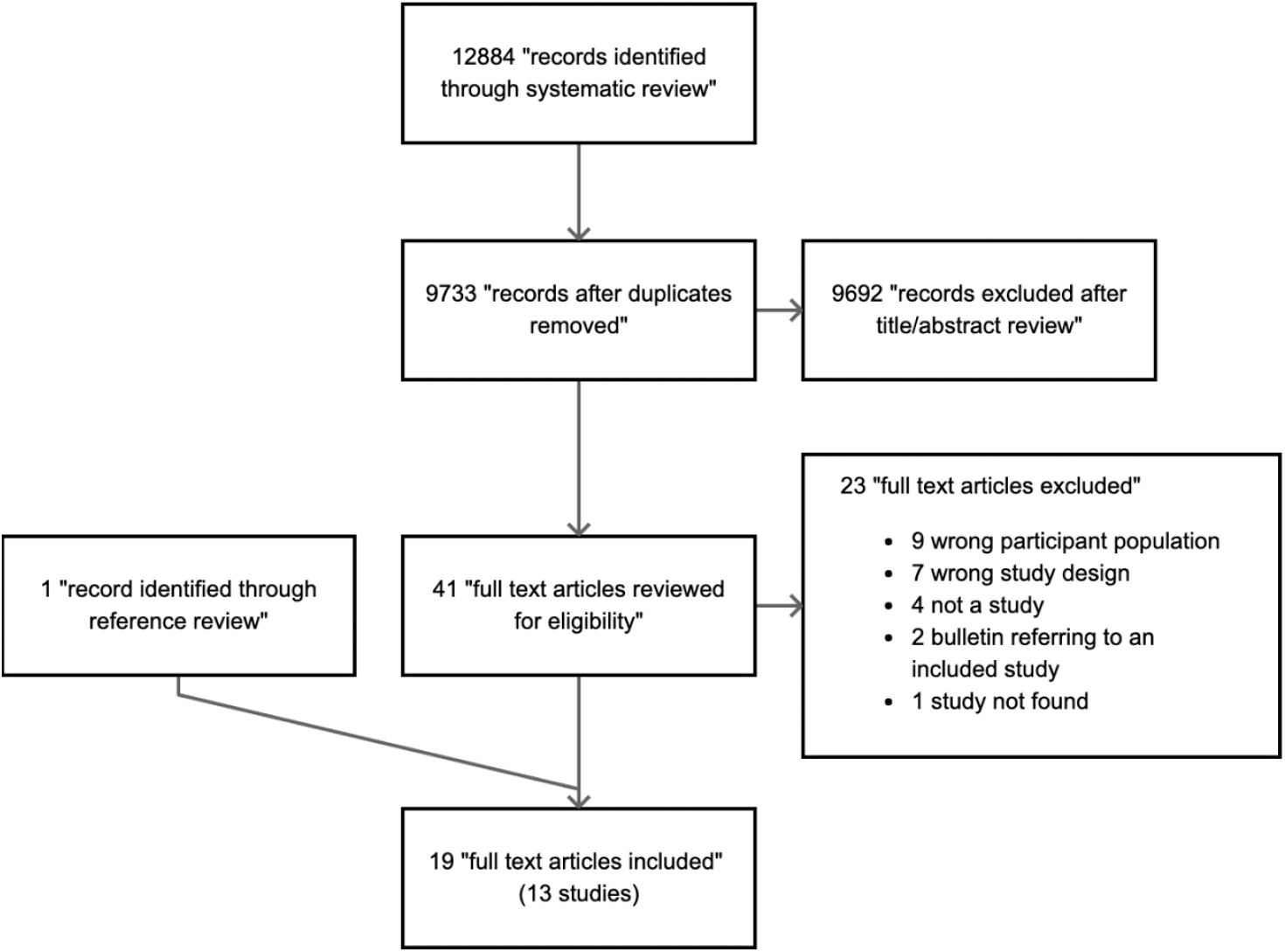
Study selection flow-diagram

### Study methodology

The studies, 12 of 13 of which were randomised controlled trials, were conducted between 1955 and 2018 across multiple WHO regions (African, European, South-East Asia, and Western Pacific) and had similar eligibility criteria (Table 1 and Supplementary Table S3). Six studies recruited participants using ACF, with the remainder recruiting participants who had presented independently to healthcare services (e.g. PCF). There was heterogeneity in the number of negative sputum cultures required at baseline (ranging from 1 to 6 samples), with only Turkova *et al* using molecular tests to define participants as microbiologically negative. There was heterogeneity in imaging modality used (e.g., plain CXR, miniature film) and methodology for CXR abnormality classification, with only two studies using predefined reporting criterion (Supplementary Table S4). Most studies compared treatment for TB against placebo or observation, with three of the more recent studies comparing different TB regimens. There was limited information on approaches to medicine administration; four studies reported on adherence.

**Table 1:** Study characteristics.

| Study | Setting and case finding methodology | Country | Baseline sputum culture-positive (excluded) | Number recruited | TB disease activity* at baseline CXR | Control Arm | Intervention Arm | Primary outcome |
| --- | --- | --- | --- | --- | --- | --- | --- | --- |
| Clayson (1963) | Not reported (ACF) | Scotland | 22/219 (10%) | 219 | Not reported | No treatment | 6HPAS** | Radiological or bacteriological diagnosis of TB at 5 years |
| Frimodt-Moller (1965) | Community CXR survey (ACF) | India | 32/468 (6.8%) | 468 | Active | Placebo | 3H (+-PAS)<br>6H (+-PAS)<br>12H (+-PAS) | Bacteriological diagnosis of TB at 4 years |
| Pamra (1971) | Community CXR survey (ACF) | India | 22/424 (5.2%) | 424 | Mix (not reported) | Placebo | 12H | Radiological or bacteriological diagnosis of TB at 6 years |
| Aneja (1979) | Symptomatic patients presenting to care (PCF) | India | 144/457 (31.5%) | 457 | Mix (75% active) | Placebo | 12HTh | Radiological or bacteriological diagnosis of TB at 1 year |
| Thompson (1982) | Annual CXR surveillance (ACF) | Multiple (Europe) | Not reported | 27830 | Inactive | Placebo | 3H<br>6H<br>12H | Bacteriological diagnosis of TB at 5 years |
| Anon (1984) | Adults presenting to care (PCF) | Hong Kong | 364/1212 (30%) | 1212 | Active | No treatment | 2SHRZ<br>3SHRZ<br>3SPH / 9S<sub>2</sub>H<sub>2</sub> | Clinical, radiological, or bacteriological diagnosis of TB at 5 years |
| Anastasatu (1985) | Hospital setting (not reported) | Romania | 230/634 (36.3%) | 634 | Active | No treatment | 3H<sub>2</sub>S<sub>2</sub>E<sub>2</sub><br>3H<sub>2</sub>S<sub>2</sub>Z<sub>2</sub><br>3R<sub>2</sub>H<sub>2</sub>S<sub>2</sub>Z<sub>2</sub> | Bacteriological diagnosis of TB at 2 years |
| Cowie (1985) | CXR surveillance in a goldmine (ACF) | South Africa | Not reported | 402 | Active | No treatment | 3RHZE | Bacteriological or histological diagnosis of TB at 5 years |
| Anon (1989) | Adults presenting to care (PCF) | Hong Kong | 592/1959 (30.2%) | 1959 | Active | No control arm | 3SHRZ<br>3S<sub>3</sub>H<sub>3</sub>R<sub>3</sub>Z<sub>3</sub><br>4S<sub>3</sub>H<sub>3</sub>R<sub>3</sub>Z<sub>3</sub> | Clinical, radiological, or bacteriological diagnosis of TB at 5 years |
| Norregaard (1990) | Adults presenting to care (PCF) | Denmark | 22/72 (30.6%) | 72 | Active | No treatment | 3RHE / 6HE | Bacteriological diagnosis of TB during follow up of at least 3 years |
| Teo (2002) | Adults presenting to care (PCF) | Singapore | 113/314 (36%) | 314 | Active | No control arm | 2HRZ / 2HR<br>2HRZ / 2H<sub>3</sub>R<sub>3</sub> | Clinical, radiological, or bacteriological diagnosis of TB at 5 years |
| Ohmori (2002) | Annual CXR surveillance (ACF) | Japan | 0/29 (0%) | 29 | Inactive | No treatment | 6H | Clinical, radiological, or bacteriological diagnosis of TB at 5 years |
| Turkova (2022) | Symptomatic children presenting to care (PCF) | Uganda, Zambia, South Africa, India | 165/1204 (14%)*** | 1204 | Active | No control arm | 2HRZ(E) / 2HR<br>2HRZ(E) / 4HR | Unfavourable status (treatment failure, treatment extension, TB recurrence, loss to follow-up, death) at 72 weeks |

\*Disease activity (active | inactive | mix) as determined by study radiologist. Inactive changes typically described as fibrotic or calcified only.
\*\*Could be extended per clinician judgement
\*\*\*Culture or Xpert MTB/RIF positive
RCT = Randomised controlled trial, ACF = Active Case Finding, PCF = Passive Case Finding, H = Isoniazid, PAS = p-aminosalicylic acid, Th = thioacetone, S = streptomycin, R = rifampicin, Z = pyrazinamide, E = ethambutol. Numbers prior to drugs denote number of months the regimen is given for. Numbers in subscript after a drug denote how many days a week it is administered on if not daily.

Apart from Turkova *et al*, few studies explicitly stated the primary outcome; however, where available, this was a diagnosis of active pulmonary TB by end of follow-up, which was at least five years for most studies. Five studies required bacteriological confirmation to meet their primary outcome of active TB, with the remainder permitting diagnosis determined by clinical assessment or progressive CXR changes. Respiratory sampling was predominantly from sputa, but some studies permitted supplementary use of laryngeal swabs or gastric lavage.

### Baseline characteristics of study participants

Most studies included more men than women. Turkova *et al* studied only children under 16 years old, whilst the rest studied adults. Although nine studies recruited participants over 15 years old, the study-level median age was generally not reported nor calculable (Supplementary Table S5). There was minimal information on symptomatology at baseline. For studies recruiting only those with active CXR changes, there appeared to be a correlation between case-finding methodology and rate of sputum culture positivity at baseline, with higher rates of culture positivity in studies using PCF (30-36%) compared to ACF (5-7%); there was no impact from the number of sputum samples collected at baseline. The reporting of baseline CXR changes varied: four studies reported on the presence of bilateral changes in keeping with TB (range 6-26%) and five reported on the presence of cavities (range 1-24%). Most studies described the CXR changes as “active”, except for Ohmori *et al* and Thompson *et al*, which required stable, fibrotic changes (e.g., “inactive” TB) as an inclusion criteria. Four studies radiographically monitored participants for up to twelve months prior to recruitment to ensure changes were not progressing. Apart from Turkova *et al*, no study clearly reported on the proportion of participants with previous TB, nor participant HIV status, although the majority were conducted prior to, or early in, the global HIV epidemic.[17] Only Ohmori *et al* and Norregaard *et al* reported on the background incidence rate of TB and most studies were conducted before TB incidence data was routinely available from the WHO.

Several treatments were evaluated and after qualitative analysis these were grouped into two categories: isoniazid-based, and multi-drug. Six studies, including the oldest five (1963 to 1982) used isoniazid-based regimens: isoniazid with or without a weak partner agent (e.g., thioacetazone, para-aminosalicylic acid) given for 3-12 months, akin to regimens used for latent TB infection. The remaining seven studies used multi-drug regimens: three or more potent agents including isoniazid and rifampicin and/or streptomycin, mostly administered for 2-3 months. Where reported, dosages of isoniazid and rifampicin were similar across trials.

### Quality assessment

Overall, the quality of included studies was low, with the oldest studies assessed as at high risk of bias (Figure 2). In the older studies reporting of study methodology was not standardised. It was often uncertain whether there had been deviation from the intended intervention, or how the randomisation had been administered, and it was frequently not possible to assess the primary outcome at multiple time points or to assess secondary outcomes (e.g., medication adherence, toxicity, clinical or radiological diagnosis of TB). Only Aneja *et al* reported using a double-blinded placebo approach in their trial. The funnel plot was asymmetrical, implying presence of publication bias (Supplementary Figure S6).

**Figure 2:**
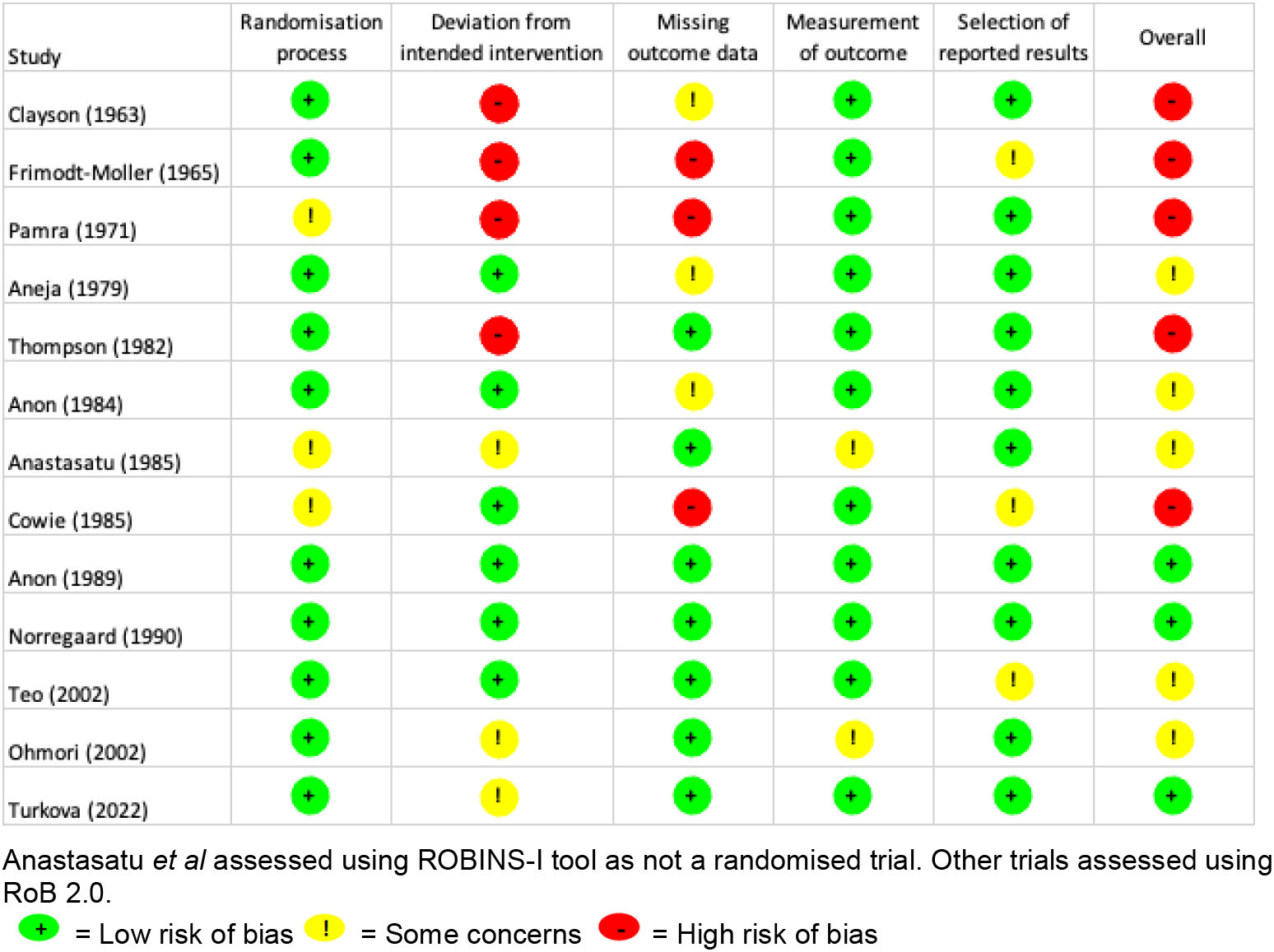
Quality assessment of included studies

### Primary outcome

Seven studies were excluded from the meta-analysis due to insufficient sampling and reporting of culture outcome data (Clayson *et al*), absence of an inactive comparator (Anon (1989), Teo *et al*, Turkova *et al*, Anastasatu *et al*), and for only including participants with solely fibrotic CXR changes (Thompson *et al*, Ohmori *et al*) (Table 2). For the remaining six studies, pooled analysis found that progression to bacteriological-confirmed pulmonary TB was significantly reduced with any treatment compared to no treatment (risk ratio [RR] 0.27, 95% CI 0.13 – 0.56, I^2^ = 90.4%) (Figure 3).

**Table 2:** Study Results*

| Study | Follow up time (years) | Control Arm |  | Intervention Arm(s) |  | Unadjusted RR (95% CI) |
| --- | --- | --- | --- | --- | --- | --- |
|  |  | Control | Risk of incident TB n/N (%) | Intervention | Risk of incident TB n/N (%) |  |
| Studies included in pooled analysis |  |  |  |  |  |  |
| Frimodt-Moller (1965) | Not reported | Placebo | 26/86 (30.2%) | 3H (+-PAS) | 28/89 (31.5%) | 1.04 (0.55 – 1.96) |
|  |  |  |  | 6H (+-PAS) | 30/83 (36.1%) | 1.20 (0.64 – 2.23) |
|  |  |  |  | 12H (+-PAS) | 17/91 (18.7%) | 0.62 (0.31 – 1.25) |
| Pamra (1971) | 6 | Placebo | 57/178 (32.0%) | 12H | 10/139 (7.2%) | 0.22 (0.12 – 0.42) |
| Aneja (1979) | 1 | Placebo | 21/110 (19.1%) | 12HTh | 11/103 (10.7%) | 0.56 (0.28 – 1.10) |
| Anon (1984) | 5 | No treatment | 71/173 (39.9%) | 2SHRZ | 10/161 (6.2%) | 0.15 (0.08 – 0.30) |
|  |  |  |  | 3SHRZ | 5/161 (3.1%) | 0.08 (0.03 – 0.19) |
|  |  |  |  | 3SPH/9S<sub>2</sub>H<sub>2</sub> | 1/160 (0.6%) | 0.02 (0.00 – 0.11) |
| Cowie (1985) | 5 | No treatment | 88/152 (57.9%) | 3HRZE | 30/250 (1.2%) | 0.21 (0.14 – 0.30) |
| Norregaard (1990) | 3 | No treatment | 8/28 (28.6%) | 3HRE/6HE | 0/22 (0%) | 0.07 (0.00 – 1.23) |
| Studies not included in pooled analysis |  |  |  |  |  |  |
| Thompson (1982) | 5 | Placebo | 97/6990 (1.4%) | 3H | 76/6956 (1.1%) | 0.79 (0.52 – 1.18) |
|  |  |  |  | 6H | 34/6965 (0.5%) | 0.35 (0.22 – 0.57) |
|  |  |  |  | 12H | 24/6919 (0.3%) | 0.25 (0.15 – 0.42) |
| Ohmori (2002) | 2.5 | No treatment | 0/15 (0%) | 6H | 0/14 (0%) | 1.07 (0.02 – 50.43) |
| Anon (1989) | 5 | n/a | n/a | 3SHRZ | 10/389 (2.6%) | n/a** |
|  |  |  |  | 3S<sub>3</sub>H<sub>3</sub>R<sub>3</sub>Z<sub>3</sub> | 10/370 (2.7%) | n/a** |
|  |  |  |  | 4S<sub>3</sub>H<sub>3</sub>R<sub>3</sub>Z<sub>3</sub> | 4/359 (1.1%) | n/a** |
| Teo (2002) | 5 | n/a | n/a | 2HRZ/2HR | 0/99 (0%) | n/a** |
|  |  |  |  | 2HRZ/2H<sub>3</sub>R<sub>3</sub> | 1/102 (1.0%) | n/a** |
| Turkova (2022) | 1.5 | n/a | n/a | 2HRZ(E)/2HR | 1/332 (0.3%) | n/a** |
|  |  |  |  | 2HRZ(E)/4HR | 0/335 (0%) | n/a** |
| Anastasatu (1985) | 2 | Adjacent cohort*** | 6/143 (4.2%) | 3H<sub>3</sub>S<sub>3</sub>E<sub>3</sub> | 5/68 (7.4%) | n/a** |
|  |  |  |  | 3H<sub>2</sub>S<sub>2</sub>Z<sub>2</sub> | 4/60 (6.7%) | n/a** |
|  |  |  |  | 3R<sub>2</sub>H<sub>2</sub>S<sub>2</sub>Z<sub>2</sub> | 2/76 (2.6%) | n/a** |

\*Clayson (1963) not included as no sputum culture data available
\*\*Not applicable due to study design
\*\*\*This arm was not a randomised comparison, but rather lower-risk participants selected for observation without treatment

**Figure 3:**
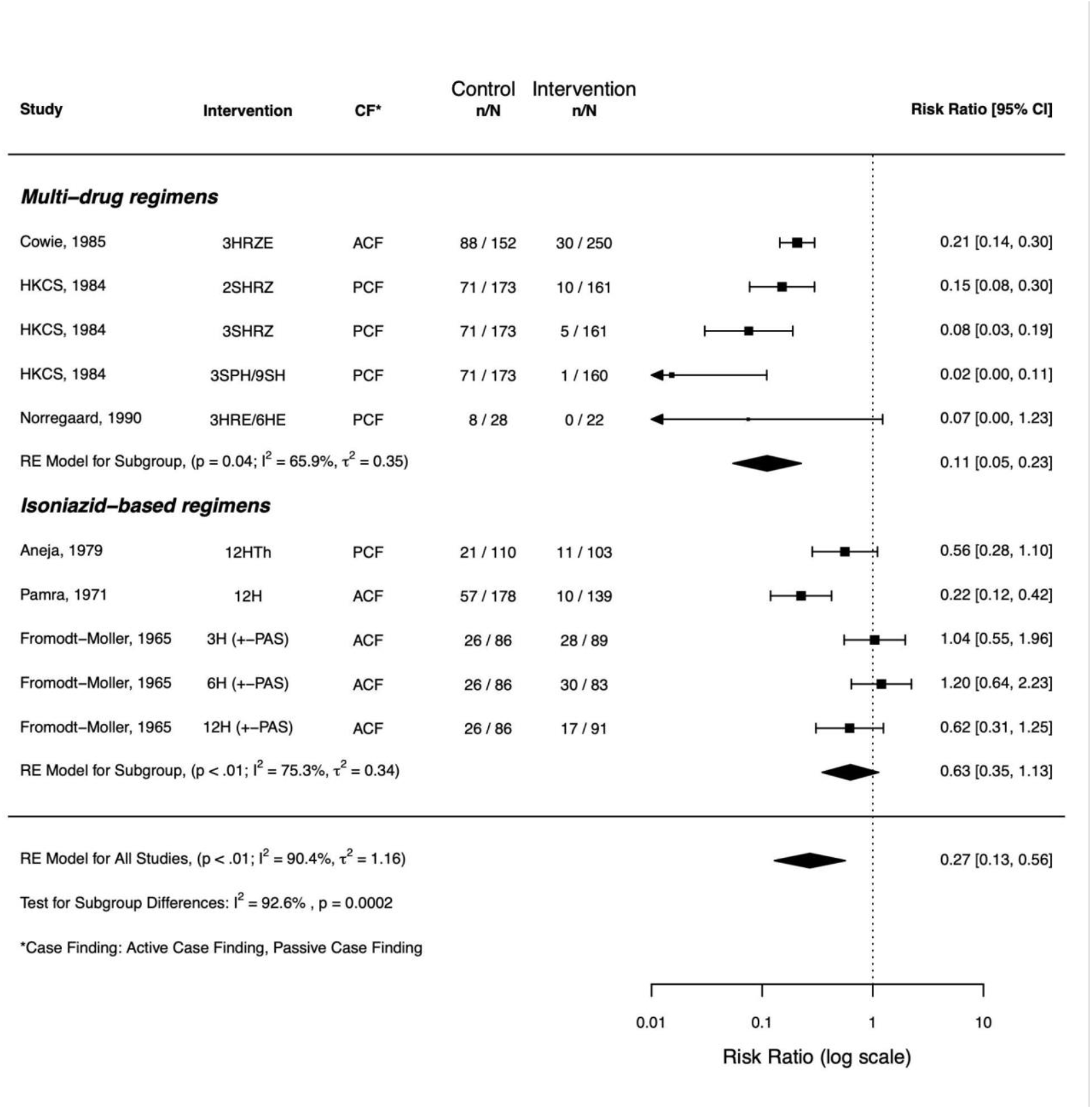
Forest plot for results against regimen.

Subgroup analysis by multi-drug vs isoniazid-based regimens found a significant difference in treatment effect (p<0.001) and reduced proportion of variability due to heterogeneity (I^2^ = 65.9% and 75.3% respectively). Multi-drug regimens conferred greater risk reduction (RR 0.11, 95% CI 0.05-0.23) in comparison to isoniazid-based regimens which showed no conclusive benefit (RR 0.63, 95% CI 0.35-1.13) (Figure 3). This effect remained unchanged in a sensitivity analysis using only regimens containing rifampicin (RR 0.15, 95% CI 0.09-0.25). The NNT for multi-drug regimens based on the pooled analysis was 2.5 (95% CI 2.2 - 3.0). Other subgroup analyses were statistically significant, with the intervention arm yielding a larger risk reduction in studies with PCF methodology compared to ACF (p=0.037), and where more than three respiratory samples were sent at baseline compared to fewer than three (p=0.018) (Supplementary Figures S7-8). However, on review of the data and degree of impact on heterogeneity, these findings appeared to be confounded by regimen type. There was insufficient data to support other subgroup analyses (HIV status, radiographic change at baseline).

Sensitivity analyses, firstly excluding studies with high risk of bias (leaving 3 studies), and where primary outcome included TB diagnosis by clinical/radiological progression (7 studies), found comparable estimates of effect to our primary analysis (Supplementary Table S9). A network analysis was conducted including the two studies conducted in Hong Kong as they had comparable intervention arms and found that longer duration of treatment was associated with improved outcomes (Supplementary Table S10).

### Secondary outcomes

Although we had planned to explore other markers of TB disease progression or regression during follow-up, no studies presented data on clinical change over time and only Clayson *et al* reported on radiographic change over time. Reporting on rates of loss to follow-up was variable, and for studies where it was clearly presented it ranged widely (0 to 17.5%). Three studies reported on adherence, with two finding a negative correlation with duration of therapy - the percentage of people taking most of their medication was higher in those allocated to 6-months treatment compared to 12-months: 87% vs 68% in Thompson *et al*, and 63% vs 25% in Frimodt-Moller *et al*.

Reporting of safety outcomes was limited and inconsistent. Two studies based in Hong Kong reported between 2-6% of participants had at least one drug stopped, although this was inclusive of participants with culture-positive TB. Cowie *et al* reported no reactions in 250 patients receiving three months of HRZE (isoniazid, rifampicin, pyrazinamide, and ethambutol), and Clayson *et al* reported 3/94 participants experiencing severe reactions whilst receiving a six-month regimen of H-PAS (isoniazid and para-aminosalicyclic acid). Drug resistance was described in seven studies, however there was substantial variation in reporting, no description of laboratory methodology, and testing was often done on a small subset. For most studies using multi-drug regimens, the rates of resistance were low (0-2%). Two studies using isoniazid-based regimens reported high rates of resistance (15-47%), but it was not possible to ascertain if there was a difference between the intervention and control arms. No study reported any patient-centred outcomes (e.g., quality of life, catastrophic costs) or recorded patient perspectives of TB treatment. Turkova *et al* reported a cost-effectiveness analysis finding reduced health care costs for those given a shorter regimen.

## DISCUSSION

This systematic review identified 13 trials in adults and children with CXR features of TB and negative sputum cultures, conducted between 1955 and 2018 in a variety of settings. Meta-analysis of six studies showed that treatment can reduce the risk of progression from culture-negative to culture-positive pulmonary TB (RR 0.27, 95% CI 0.13 - 0.56). The risk of progression to culture-positive TB for individuals with active CXR changes without treatment was substantial (19-58%). For those treated with a multi-drug regimen, the rate was substantially and significantly lower than compared to placebo or observation (RR 0.11, 95% CI 0.05 - 0.23), and although there was an impact from duration, regimens of 2-3 months appeared to offer excellent results. Although we acknowledge limitations of pooled NNT calculations, a NNT of 2.5 is substantially lower than estimates for treatment of latent tuberculosis infection (NNT of 100).[18] Studies using isoniazid-based regimens did not show the same impact of treatment (0.63, 95% CI 0.35-1.13).

Current guidelines on the management of this group of patients is variable and non-specific. A trial of antibiotics is frequently recommended followed by clinical discretion,[19] though recent evidence refutes the evidence base for this approach highlighting the potential harm of delayed diagnosis and treatment.[20] The Centres for Disease Control and Prevention / Infectious Diseases Society of America guidelines support the use of a four-month regimen based on the findings from the Hong Kong studies,[21] however, implementation of this in practice is low. Most existing guidelines have not incorporated evidence from the studies included in this review, which is unsurprising as the regimens evaluated are largely outdated, and the described phenotype (CXR positive, culture-negative) is not officially recognised. WHO guidelines for management of latent TB infection suggest that if an individual with CXR changes has active TB excluded bacteriologically, preventive therapy could be considered. The results of this review provide evidence to suggest that IPT (isoniazid preventive therapy), particularly if given for only six months, would not be appropriate for patients with CXR changes suggestive of active TB, even if sputum culture is negative. There is currently little evidence to support the use of other preventive regimens for this group; however, experience of using four months of rifampicin and isoniazid in an observational cohort of 414 patients found very low disease progression (1.2% culture-positive over 60 months), although this is yet to be evaluated in a clinical trial.[22] All included studies were conducted prior to or early on in the global HIV epidemic, with no discussion of recurrent TB infections and its relevance to chronic CXR changes, and before the introduction of molecular diagnostics. False positive sputum PCR results (PCR positive, culture-negative) have been described several years after successful treatment completion and their relevance is poorly understood.[23]

Our findings add weight to the idea that patients in the middle of the TB disease spectrum are distinct to patients with culture-positive disease or latent TB and should be managed differently.[8] Given the rapid expansion of CXR-based screening programmes globally, this group of patients will be increasingly identified. This review highlights the need for development of clinical trials aiming to identify appropriate diagnostic and treatment options. Additionally, our review has broader relevance for the development of trials hoping to incorporate novel biomarkers (e.g., blood transcriptomics) and advanced radiological methods (e.g., positron emission tomography / computed tomography) which may better identify patients at increased risk of disease progression to culture-positive disease.[24]

This systematic review has limitations. The number of identified studies was small, some of which had low participation, and there was significant between-study heterogeneity. A variety of regimens and durations were used (many of which contained drugs now rarely used). However, we were able to broadly categorise treatment into isoniazid-based and multi-drug, which provided valuable insights. We were not able to explore differences in treatment effects by symptomatology due to limited reporting and confounding factors (case-finding methodology, regimen composition). We recognise that variation in follow-up and missing baseline, outcome, and safety data could have introduced reporting bias, though reassuringly sensitivity analyses showed no change in treatment effect. Missing or incomparable data due to heterogeneity of study methodology meant several pre-defined sub-group analyses were not possible. Most studies were conducted over 25 years ago and there have been advances in the diagnostic approach to TB with respect to imaging (e.g., digital x-ray, computer-aided-detection) and microbiological techniques (e.g., molecular diagnostics, liquid culture), likely improving sensitivity and specificity. Furthermore, laboratories would have been less well equipped by modern standards, with limited or no quality assurance, potentially leading to misclassification of the primary outcome by either under or over-reporting. Laboratory capability to observe for acquired drug resistance was also limited. The lack of presented data on specific participant comorbidities (e.g., diabetes, HIV) limits the applicability of findings to these groups. Six studies excluded participants with prior TB history, increasing the probability that CXR changes were due to active disease, but reducing the generalisability of our results.

Rifapentine-based regimens compared to rifampicin-based regimens have recently been shown to reduce the required treatment duration for culture-positive disease and latent TB infection to four months and one month respectively,[25, 26] however, none of the regimens evaluated in this review contained rifapentine. It is therefore possible that treatment regimens shorter than four months might be effective for those with negative cultures.

Despite limitations, this review finds that abbreviated, multi-drug regimens for culture-negative pulmonary TB can prevent progression to culture-positive disease. Isoniazid, particularly if used for only six months, appears unlikely to prevent progression. This study population is becoming increasingly relevant as CXR-based screening expands and diagnostic tests with the potential to identify incipient TB become available. Modern clinical trials are necessary in order to establish the optimal approach to treatment in this patient group as we re-define the previously accepted binary approach to TB management.

## Data Availability

All data produced in the present study are available upon reasonable request to the authors

## Acknowledgements

We thank Dr Itaru Nakamura, Associate Professor at the Department of Infection Prevention and Control, Tokyo Medical University Hospital, Japan, for his help extracting data from a relevant article published in Japanese.

We thank Dr Anna Turkova and Prof Angela Crook, at the Medical Research Council Clinical Trials Unit, University College London and her team for provision of unpublished subgroup data from the SHINE trial.

