## Supplementary Material for "Treatment for radiographically active, sputum culture-negative pulmonary tuberculosis: a systematic review and meta-analysis"

#### Title

### Supplementary Material

**Example search strategy for MEDLINE (Ovid, from 1946)**

1. Mycobacterium tuberculosis/
2. tuberculosis.ti, ab, kf
3. TB.ti, ab, kf
4. exp tuberculosis/
5. 1 or 2 or 3 or 4
6. (Abacillary or "doubtful activity").ti, ab, kf
7. (culture adj2 negative).ti, ab, kf
8. (smear adj2 negative).ti, ab, kf
9. (xray or x ray or x-ray or cxr).ti, ab, kf
10. 6 or 7 or 8 or 9
11. exp Antitubercular Agents/
12. Mycobacterium tuberculosis/de [Drug Effects]
13. Tuberculosis/dt [Drug Therapy]
14. chemotherapy, adjuvant/ or drug therapy, combination/
15. Drug Administration Schedule/
16. Tuberculosis, Pulmonary/th [Therapy]
17. antitubercul* agent*.ti, ab, kf
18. antitubercul* drug*.ti, ab, kf
19. chemotherapy.ti, ab, kf
20. exp Drug Therapy/
21. 11 or 12 or 13 or 14 or 15 or 16 or 17 or 18 or 19 or 20
22. 5 and 10 and 21
23. exp Animals/
24. Humans/
25. 23 not 24
26. 22 not 25

**Supplementary table S1: Definitions of secondary outcomes**

| **Secondary Outcome** | **Definition** |
|---|---|
| Radiologically confirmed TB | Progression of CXR changes consistent with TB during follow-up as compared to baseline, with no microbiological confirmation, and in the absence of an alternative diagnosis |
| Clinical evidence of tuberculosis | Symptoms or clinical findings leading to a diagnosis of active tuberculosis by the treating physician, without supportive microbiological or radiographic evidence |
| Safety outcomes | Patient adverse events reported during the study period from the first day of intervention, whether or not attributed to study intervention |

| | |
|---|---|
| Resistance | Documentation of laboratory confirmed resistance to one or more anti-TB drugs included in the treating regimen |
| Treatment completion | Included participants who started the intervention and completed the intended course as per study protocol |
| Loss to follow-up | Any patient who did not complete the intended follow-up period of the study, and as a result the primary outcome was unknown |

**List of included studies**


1. Clayson C, Research committee of the Scottish Thoracic Society. A controlled trial of chemotherapy in pulmonary tuberculosis of doubtful activity. *Tubercle* 1958;39(3):129-137
2. Clayson C, Research committee of the Scottish Thoracic Society. A controlled trial of chemotherapy in pulmonary tuberculosis of doubtful activity: five year follow-up. *Tubercle* 1963;44(1):39-46
3. Frimodt-Moller J, Parthasarathy R, Thomas J. Results of treatment of non-bacillary tuberculosis in a domiciliary - a preliminary report. *Tuberculosis and chest diseases worker's conference* 1960
4. Pamra SP, Mathur GP. Effects of chemoprophylaxis on minimal pulmonary tuberculosis lesions of doubtful activity. *Bull WHO* 1971;45:593-602
5. Aneja KS, Gothi GD, Rupert Samuel GE. Controlled study of the effect of specific treatment on bacteriological status of 'suspect cases'. *Ind J Tub* 1979;26(2):50-57
6. Hong Kong Chest Service/Tuberculosis Research Centre Madras/British Medical Research Council. Sputum-smear-negative pulmonary tuberculosis. Controlled trial of 3-month and 2-month regimens of chemotherapy. First report. *Lancet* 1979;1(8131):1361-1368
7. Girling DJ. Hong Kong Chest Service/Tuberculosis Research Centre Madras/British Medical Research Council study of two-month and three-month regimens for smear-negative pulmonary tuberculosis: results up to two years. *Br J Dis Chest* 1979:73:413
8. Hong Kong Chest Service/Tuberculosis Research Centre Madras/British Medical Research Council. A controlled trial of 2-month, 3-month, and 12-month regimens of chemotherapy for sputum-smear-negative pulmonary tuberculosis: the results at 30 months. *Am Rev Resp Dis* 1981;124(2):138-142
9. Hong Kong Chest Service/Tuberculosis Research Centre Madras/British Medical Research Council. A controlled trial of 2-month, 3-month, and 12-month regimens of chemotherapy for sputum-smear-negative pulmonary tuberculosis. Results at 60 months. *Am Rev Resp Dis* 1984;130(1):23-28
10. Thompson NJ, International union against tuberculosis committee on prophylaxis. Efficacy of various durations of isoniazid preventive therapy for tuberculosis: five years of follow-up in the IUAT trial. *Bull WHO* 1982;60(4):555-564
11. Chan SL. Hong Kong Chest Service/Tuberculosis Research Centre Madras/British Medical Research Council studies of smear-negative pulmonary tuberculosis. *Bull Int Un Tuberc* 1985;60(3-4):106-107
12. Hong Kong Chest Service/Tuberculosis Research Centre Madras/British Medical Research Council. A controlled trial of 3-month, 4-month, and 6-month regimens of chemotherapy for sputum-smear-negative pulmonary tuberculosis. Results at five years. *Am Rev Resp Dis* 1989;139(4):871-6
13. Anastasatu C, Bercea O, Corlan E. Controlled clinical trial on smear-negative, x-ray positive new cases, with a view to establish if and how to treat them. *Bull Int Union Tuberc Lung Dis* 1985;60:108-109
14. Cowie RL, Langton ME, Escreet BC. Diagnosis of sputum smear- and sputum culture-negative pulmonary tuberculosis. *S Afr Med J* 1985;68(12):878
15. Cowie RL, Langton ME, Escreet BC. Ultrashort-course chemotherapy for culture-negative pulmonary tuberculosis – a qualified success. *S Afr Med J* 1985;68(12):879-880
16. Norregaard J, Heckscher T, Viskum K. Abacillary pulmonary tuberculosis. *Tubercle* 1990;71:35-38

17. Teo SK, Tan KK, Khoo TK. Four-month chemotherapy for the treatment of smear-negative pulmonary tuberculosis: results at 30 to 60 months. *Ann Acad Med Singap* 2002;31(2):175-81
18. Ohmori M, Wada M, Nishii K, et al. [Preventive therapy in middle-aged and elderly persons selected from the population-based screening by mass miniature radiography-methodological aspect and adverse reactions]. *Kekkaku* 2002;77(10):647-58.
19. Turkova A, Wills GH, Wobudeya E, et al. Shorter treatment for nonsevere tuberculosis in african and indian children. *N Engl J Med* 2022;386:911-922.

**Supplementary table S2: Excluded studies at full text review**

| Study title | Reason for exclusion |
|---|---|
| Grant. Observations on cases with minimal tuberculosis of doubtful activity with or without chemotherapy. *Zeitschrift fur tuberkulose und erkrankungen der thoraxorgane* 1966;125(3):198-201 | Inappropriate study design |
| Ade S, Harries AD, Trebucq A, et al. National profile and treatment outcomes of adult smear-negative pulmonary TB patients in Benin. *Trans R Soc Trop Med Hyg* 2013;107(12):783-8 | Inappropriate study design |
| Ormerod LP, Green RM, Horsfield N. Outcome of the treatment of culture negative tuberculosis (respiratory and non-respiratory): Blackburn 1996-2000. *J Infect* 2002;45(2):88-9 | Inappropriate study design |
| Ormerod LP, McCarthy OR, Rudd RM, et al. Short-course chemotherapy for tuberculosis pleural effusion and culture-negative pulmonary tuberculosis. *Tuber Lung Dis* 1995;76(1):25-7 | Inappropriate study design |
| Dutt AK, Moers D, Stead WW. Smear-negative and culture-negative pulmonary tuberculosis - 4-month short-course chemotherapy. *Am Rev Resp Dis* 1989;139(4):867-870 | Inappropriate study design |
| Ferebee SH, Mount FW, Murray FJ, et al. A controlled trial of isoniazid prophylaxis in mental institutions. *Am Rev Respir Dis* 1963;88:161-75 | Inappropriate study design |
| Krishnaswami KV. Fate of abacillary pulmonary tuberculosis cases. *Indian J Chest Dis Allied Sci* 1976;18(3):133-140 | Inappropriate study design |
| Rubin EH, Cheifetz I, Katzev H, et al. Isoniazid in pulmonary tuberculosis, with special reference to x-ray findings after four and six months' treatment. *Med Clin North Am* 1953;1:885-901 | Irrelevant patient population |
| Chan SL, Wong PC, Tam CM. 4-, 5- and 6-month regimens containing isoniazid, rifampicin, pyrazinamide and streptomycin for treatment of pulmonary tuberculosis under program conditions of Hong Kong. *Tubercle Lung Dis* 1994;75(4):245-50 | Irrelevant patient population |
| Zierski M, Bek E, Long MW, et al. Short-course (6 month) cooperative tuberculosis study in Poland: results 18 months after completion of treatment. *Am Rev Resp Dis* 1980;122(6):879-89 | Irrelevant patient population |
| Hong Kong Chest Service/Tuberculosis Research Centre Madras/British Medical Research Council. A controlled clinical comparison of 6 and 8 months of antituberculosis chemotherapy in the treatment of patients with silicotuberculosis in Hong Kong. *Am Rev Resp Dis* 1991;143(2):262-7 | Irrelevant patient population |
| Yanuar M, Nawas A, Hudoyo A, et al. Evaluation of diagnosis and treatment of pulmonary tuberculosis smear acid fast bacilli negative in jakarta respiratory centre (JRC). *Respirology* 2010;15(s2):62 | Irrelevant patient population |

| | |
|---|---|
| Chakraborty H. Treatment of MMR detected shadows of doubtful activity with INH alone. *J Ind Med Assoc.* 1972;58(5):162-4 | Irrelevant patient population |
| Baba H, Shinkai A, Azuma Y. Controlled clinical trial of three 6 month regimens of chemotherapy for pulmonary tuberculosis (Report 2). Results at one year after completing chemotherapy. *Kekkaku* 1979;54(1):29-36 | Irrelevant patient population |
| Ovchinnikova IE, Starshinova AA, Dovgaliuk IF. [Optimization of chemotherapy regimens in children with primary pulmonary tuberculosis]. *Problemy tuberkuleza I boleznei legkikh* 2009;(1):36-40 | Irrelevant patient population |
| Horwitz O, Payne PG, Wilbek E. Epidemiological basis of tuberculosis eradication. 4.The Isoniazid trial in Greenland. *Bull WHO* 1966;35(4):509-26 | Irrelevant patient population |

**Supplementary table S3: Study design and eligibility criteria**

| Study | Number of negative sputum cultures | Chest x-ray criteria | Clinical criteria | Other criteria | Exclusion criteria |
|---|---|---|---|---|---|
| Clayson (1963) | 1 | "Radiographic abnormality considered to be TB… …and no cavitation" | Physician prepared to not treat | Age > 15 years old<br>European | Extra-pulmonary TB<br>Pregnancy<br>Diabetes<br>Known to TB service<br>Disease obviously due to primary infection |
| Frimodt-Moller (1965) | Not reported | "X-ray lesions consistent with pulmonary TB… …which are not calcified" | None | None | On TB treatment |
| Pamra (1971) | 2 | "presumably tuberculous pulmonary lesions"<br>"radiological stability over at least three months" | Asymptomatic | None | Previous TB treatment<br>Diabetes<br>Extra-pulmonary TB<br>Non-tuberculosis mycobacteria disease |
| Aneja (1979) | 1 | "Abnormal shadow on photofluorogram" | Well enough to attend outpatients | Aged > 12 years old | Previous treatment for TB lasting >2 weeks |

| | | | | | |
|---|---|---|---|---|---|
| Thompson (1982) | 2 | "Fibrotic lung lesions of probable tuberculous origin, on chest x-ray stable for 12 months… … and not limited to solitary calcifications or pleural thickening" | None | Tuberculin skin test positive >6mm | Previous TB treatment |
| Anon (1984) | 5 | "Radiographically active pulmonary TB" | None | Aged 15-75 years old | Previous TB treatment |
| Anastasatu (1985) | Not reported | "Pulmonary lesions suggestive of TB" | None | None | None |
| Cowie (1985) | 2 | "New or enlarging apical lung lesions, persistent over two months" | None | Tuberculin skin test (5TU) >9mm after 72 hours | None |
| Anon (1989) | 4 | "Radiologically active TB" | None | Aged 15-75 years old | Previous TB treatment |
| Norregaard (1990) | 6 | "x-ray shadows consistent with active TB" | None | None | None |
| Teo (2002) | 4 | "Abnormal chest x-ray compatible with a diagnosis of pulmonary TB" | Respiratory symptoms | None | Previous TB<br>Mental health disorder<br>Alcohol or substance use disorder<br>Pregnancy |
| Ohmori (2002) | 3 | "fibrous lesion compatible with healed TB stable for 1 years" | None | None | Severe "disease"<br>Severe liver dysfunction<br>Previous TB |
| Turkova (2022) | 2 | "confined to one lobe with no cavities, no signs of military TB, no complex pleural effusion…" | Symptomatic, non-severe TB | Aged <16 years old | Suspected DR-TB<br>Pregnancy |

**Supplementary table S4: Outcome assessments**

| Study | Microbiological assessment | | Radiographic assessment | |
|---|---|---|---|---|
| | Method | Frequency | Method | Frequency |

| | | | | |
|---|---|---|---|---|
| Clayson (1963) | Sputum or gastric lavage or laryngeal swab smear and culture | Every 3 months for two years, then every 12 months for three years. Additional testing if clinical or radiographic deterioration | Miniature film reported by a central blinded panel on two separate occasions | Every 3 months for two years, then every 12 months for three years. Additional testing if clinical deterioration |
| Frimodt-Moller (1965) | Sputum and laryngeal swab smear and culture | Every 3 months for four years | Chest x-ray reported by two independent readers and re-examined by a blinded reader | Every 3 months for four years |
| Pamra (1971) | Sputum and laryngeal swab culture | Every 3 months for one year, then every 6 months for five years | Miniature film reported by two experienced readers with a third reader if disagreement | Every 3 months for one year, then every 6 months for five years |
| Aneja (1979) | Sputum smear and culture | Every 2 months for six months, then every 3 months for six months | Photofluorogram reported by a medical officer and study readers, with disagreement resolved by an umpire | Every 2 months for six months, then every 3 months for six months. |
| Thompson (1982) | Sputum culture | Every 12 months for five years | Chest x-ray reported by a single reader | Every 12 months for five years |
| Anon (1984) | Sputum smear and culture | Every 1 month for one year, then every 3 months for four years | Chest x-ray reported at “physician meetings” | Every 1 month for three months, then every 2 months for nine months, then every 12 months for four years |
| Anastasatu (1985) | Sputum smear and culture | Frequency not reported | Chest x-ray. Procedure for description not reported | Frequency not reported |
| Cowie (1985) | Sputum smear and culture | If clinical or radiographic progression | Chest x-ray. Procedure for description not reported | Every 3 months for three years, then every 6 months for two years |
| Anon (1989) | Sputum smear and culture | Every 1 month for five years | PA chest x-ray reported at “physician meetings” with an independent blinded assessment | Every 6 months for one year, then every 12 months for four years |
| Norregaard (1990) | Sputum smear and culture (gastric lavage if unable to expectorate) | Every 1 month for three months, then every 2 months for six months, then every 12 months for two years | Chest x-ray reported by a “conference of consultants” | Every 1 month for three months, then every 2 months for six months, then every 12 months for two years. |
| Teo (2002) | Sputum smear and culture | Every 1 month for one year, then every 3 months for one year, then every 6 months for three years | Chest x-ray reported by a blinded radiologist | Every 4-6 months for one year, then every 12 months for four years |
| Ohmori (2002) | Not reported | n/a | Chest x-ray. Procedure for description not reported | Every 6 months for five years |

| | | | | |
|---|---|---|---|---|
| Turkova (2022) | Sputum smear, Xpert MTB/RIF assay, and culture | If previous samples positive, clinical suspicion of treatment failure, or new contact with DR-TB | Chest x-ray reported by two, independent, blinded experts. Procedure described in study protocol | Frequency not reported |

***** 70 participants did not get a baseline large film (which was used to estimate size of lesion), therefore relied on result from miniature film.

**Supplementary table S5: Participant characteristics**

| Study | Age (years old) | Gender | Symptoms at baseline | Cavitatory lung disease at baseline | Bilateral changes at baseline |
|---|---|---|---|---|---|
| Clayson (1963) | 15-20 (21)<br>21-30 (47)<br>31-40 (44)<br>40+ (77) | Male (110)<br>Female (79) | Not reported | n/a (excluded) | 49/189 (26%) |
| Frimodt-Moller (1965) | 15-34 (104)<br>35-44 (76)<br>45+ (169) | Male (248)<br>Female (101) | Not reported | 84/349 (24%) | 28/349 (8%) |
| Pamra (1971) | 15-24 (39)<br>25-34 (122)<br>25-44 (103)<br>45+ (53) | Male (277)<br>Female (40) | 0/317 (0%) had symptoms | Not reported | 20/317 (6%) |
| Aneja (1979) | 12-24 (37)<br>25-44 (104)<br>45+ (72) | Male (131)<br>Female (82) | 205/213 (96%) cough >2 weeks | 38/213 (18%) | Not reported |
| IUAT (1982) | Not reported | Male (14750)<br>Female (13080) | Not reported | Not reported | Not reported |
| Anon (1984) | 15-24 (300)<br>25-34 (168)<br>34-44 (88)<br>45-54 (86)<br>55+ (49) | Male (509)<br>Female (182) | “Majority with symptoms” | 7/691 (1%) | Not reported |
| Anastasatu (1985) | Not reported | Not reported | Not reported | Not reported | Not reported |

| | | | | | |
|---|---|---|---|---|---|
| Cowie (1985) | Not reported | Male (402)<br>Female (0) | Not reported | Not reported | Not reported |
| Anon (1989) | 15-24 (565)<br>25-34 (302)<br>35-44 (108)<br>45-54 (81)<br>55+ (62) | Male (724)<br>Female (394) | 1118/1118 (100%) had symptoms | 12/1118 (1%) | Not reported |
| Norregaard (1990) | 20-39 (10)<br>40-59 (17)<br>60-69 (13)<br>70+ (10) | Male (44)<br>Female (6) | 33/50 (66%) had cough | 3/50 (6%) | 6/50 (12%) |
| Teo (2002) | 15-34 (193)<br>35-54 (93)<br>55+ (28)* | Male (235)<br>Female (79) | 0/314 (0%) had symptoms | Not reported | Not reported |
| Ohmori (2002) | Not reported<br>Median: 66 | Male (15)<br>Female (14) | 0/29 (0%) had symptoms | Not reported | Not reported |
| Turkova (2022) | Not reported<br>Median: 3.5 | Male (621)<br>Female (583) | 1204/1204 (100%) had symptoms | n/a (excluded) | n/a (excluded) |

*Age details for Teo (2002) include patients excluded as culture positive

**Supplementary figure S6: Funnel plot for assessment of publication bias**

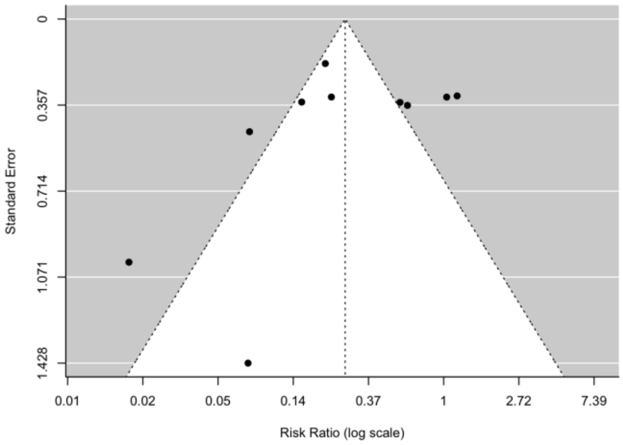


**Supplementary table S7: Forest plot of primary outcome stratified by case-finding strategy**

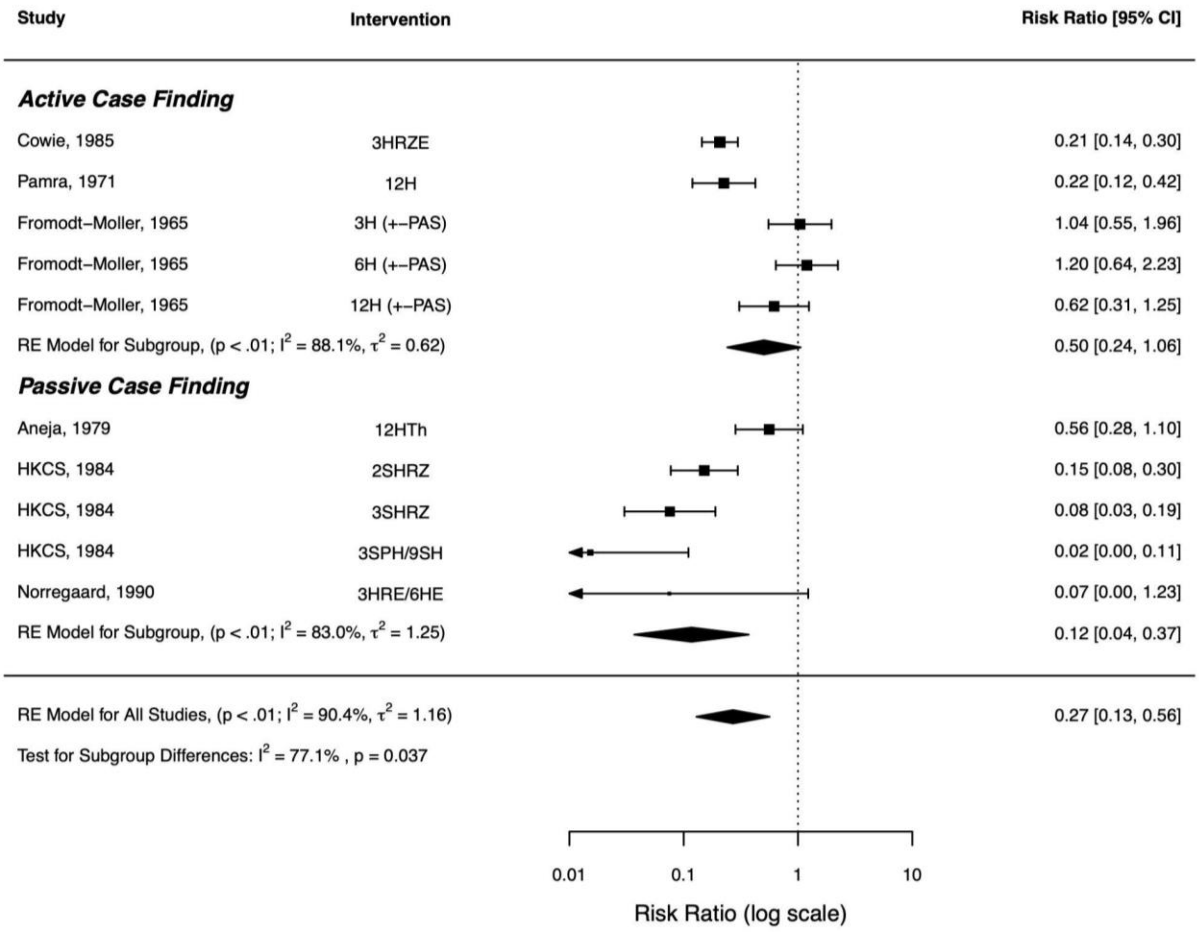


**Supplementary table S8: Forest plot of primary outcome stratified by number of baseline sputa**

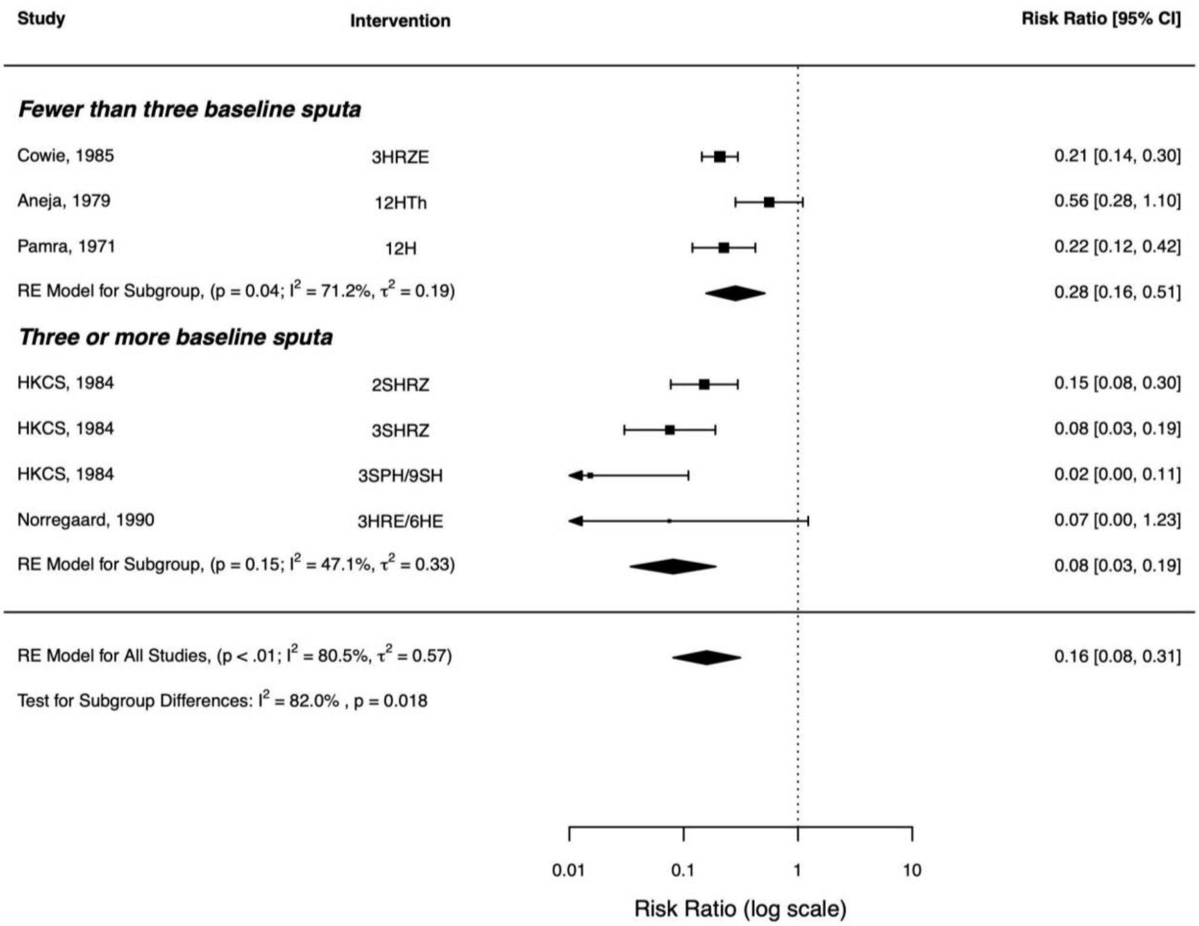


**Supplementary table S9: Sensitivity analyses**

| Analysis criteria | Number of studies | RR (95% CI) |
|---|---|---|
| Only studies with low or some risk of bias | 3 | 0.12 (0.04 - 0.37) |
| All TB diagnoses (e.g., clinical or radiographic) as the primary end-point (as opposed to only bacteriologically confirmed) | 7 | 0.32 (0.18 - 0.58) |
| Using per-protocol results | 3 | 0.14 (0.05 - 0.39) |
| Only rifampicin containing regimens | 3 | 0.15 (0.09 – 0.25) |

**Supplementary table S10: Network meta-analysis**

| Study | Intervention | Events | Group Size | Group |
|---|---|---|---|---|
| Anon (1984) | No treatment | 71 | 173 | No_trt |
| Anon (1984) | 2SHRZ | 10 | 161 | 2mth_trt |
| Anon (1984) | 3SHRZ | 5 | 161 | 3mth_trt |

| | | | | |
|---|---|---|---|---|
| Anon (1984) | $3SPH/9S_2H_2$ | 1 | 160 | 12mth_trt |
| Anon (1989) | 3SHRZ | 10 | 389 | 3mth_trt |
| Anon (1989) | $4S_3H_3R_3Z_3$ | 4 | 359 | 4mth_trt |

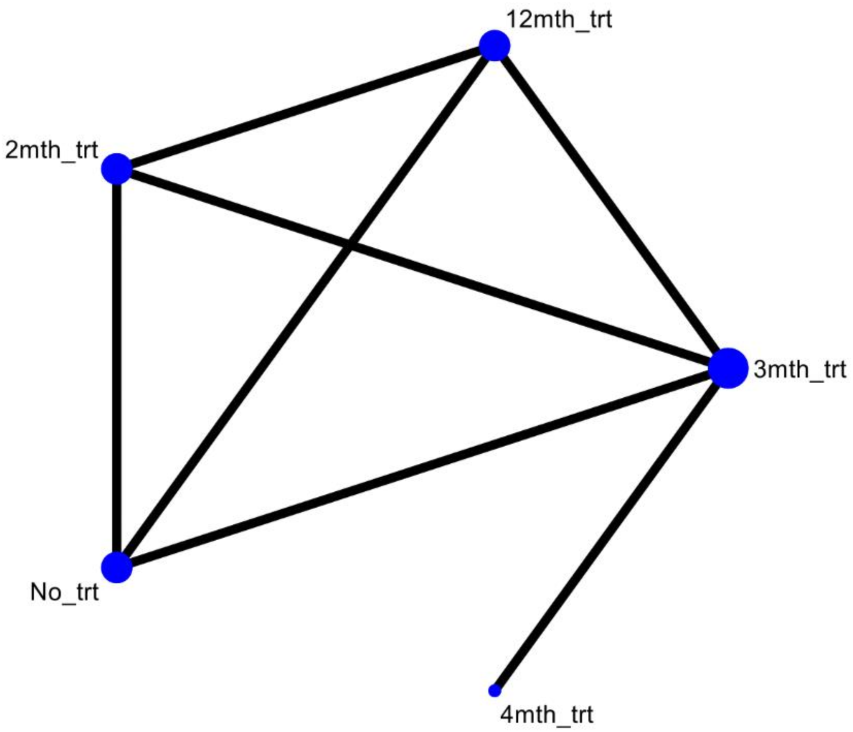


| **Rank** | **12mth_trt** | **2mth_trt** | **3mth_trt** | **4mth_trt** | **No_trt** |
|---|---|---|---|---|---|
| Best | 73.5 | 0.1 | 0.5 | 26.0 | 0.0 |
| 2nd | 19.9 | 1.8 | 13.1 | 65.1 | 0.0 |
| 3rd | 5.7 | 9.5 | 77.4 | 7.3 | 0.0 |
| 4th | 0.8 | 88.6 | 8.9 | 1.6 | 0.0 |
| Worst | 0.0 | 0.0 | 0.0 | 0.0 | 100.0 |
| **Mean Rank** | **1.3** | **3.9** | **2.9** | **1.8** | **5.0** |
| **SUCRA** | **0.9** | **0.3** | **0.5** | **0.8** | **0.0** |
